# SARS-CoV-2 receptor is co-expressed with elements of the kinin-kallikrein, renin-angiotensin and coagulation systems in alveolar cells

**DOI:** 10.1101/2020.06.02.20120634

**Authors:** Davi Sidarta-Oliveira, Carlos Poblete Jara, Adriano J. Ferruzzi, Munir S. Skaf, William H. Velander, Eliana P. Araujo, Licio A. Velloso

## Abstract

SARS-CoV-2, the pathogenic agent of COVID-19, employs angiotensin converting enzyme-2 (ACE2) as its cell entry receptor. Clinical data reveal that in severe COVID-19, SARS-CoV-2 infects the lung, leading to a frequently lethal triad of respiratory insufficiency, acute cardiovascular failure, and coagulopathy. Physiologically, ACE2 plays a role in the regulation of three systems that could potentially be involved in the pathogenesis of severe COVID-19: the kinin-kallikrein system, resulting in acute lung inflammatory edema; the renin-angiotensin system, promoting cardiovascular instability; and the coagulation system, leading to thromboembolism. Here we analyzed ~130,000 human lung single-cell transcriptomes and show that key elements of the kinin-kallikrein, renin-angiotensin and coagulation systems are co-expressed with ACE2 in alveolar cells, which could explain how changes in ACE2 promoted by SARS-CoV-2 cell entry result in the development of the three most severe clinical components of COVID-19.

## Introduction

COVID-19, the disease caused by SARS-CoV-2 infection, frequently opens with cough, fever, fatigue, and myalgia (*1*), progressing to a severe illness in up to 20% of infected patients (*2*). Severe COVID-19 is characterized by progressive dyspnea, which results from acute lung inflammatory edema leading to hypoxia (*3*). In patients surviving the initial lung inflammatory burst, a number of other complications can sum to promote a rapid and frequently lethal deterioration of health (*3-5*). Acute cardiovascular failure and coagulopathy are among the most frequent complications and could be placed alongside with acute respiratory failure (*3-7*) as components of a triad that leads to the highest death rates in COVID-19 (Fig. 1A).

**Figure 1.**
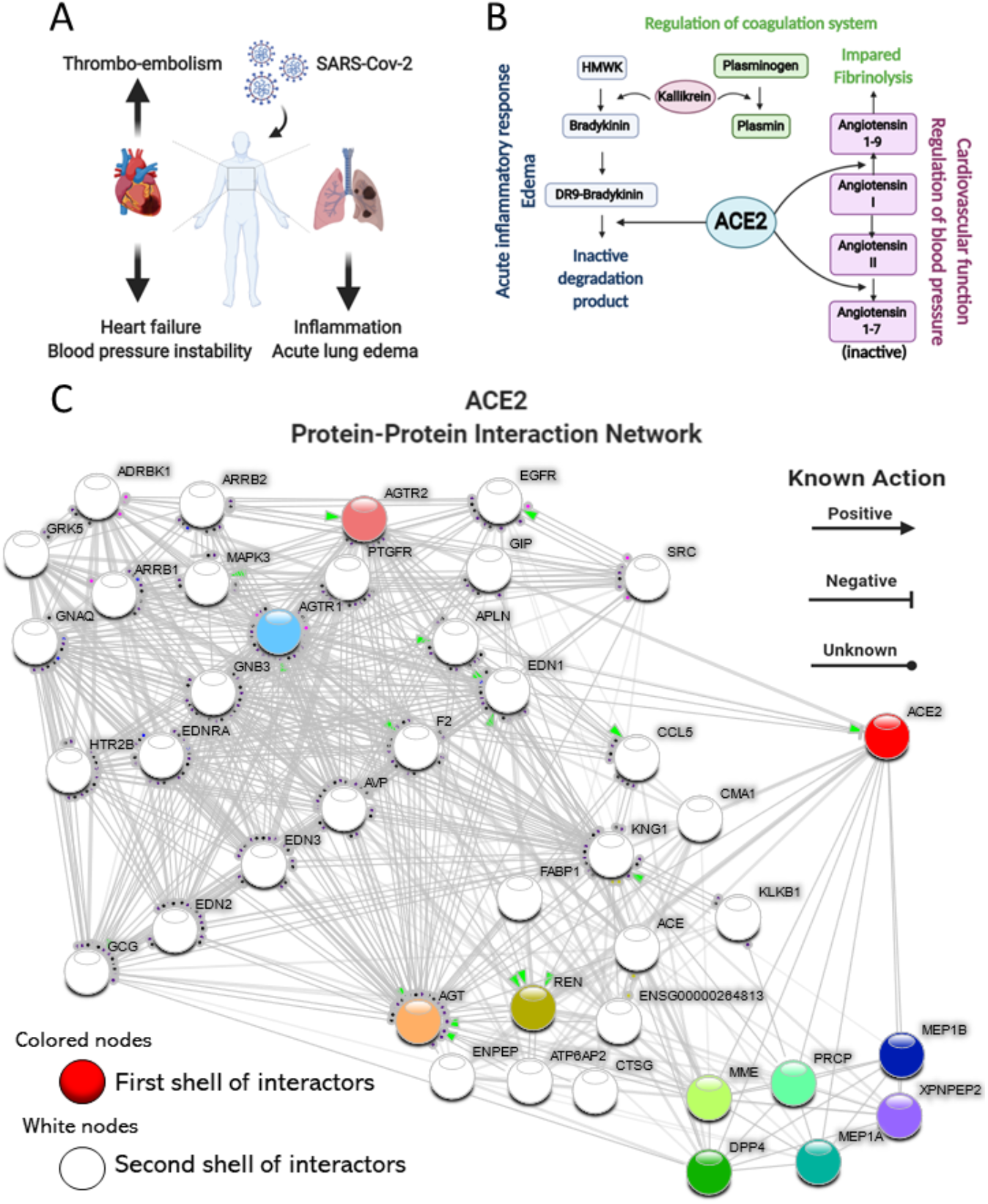
COVID-19 clinical triad and ACE2 physiological roles link COVID-19 to ACE2 dysfunction. (A) Schematic representation of the clinical triad that play central role in severe COVID-19. (B) Schematic representation of the key components of the kinin-kallikrein, renin- angiotensin and coagulation systems and their interfaces with ACE2. (C) ACE2 Protein-Protein Interaction Network. ACE2 interactome was retrieved with the data-mining toolkit string-db (https://version-11-0.string-db.org/cgi/network.pl?taskId=KwL0Kho7pBZf). First shell interactors of ACE2 were set as colored nodes, while second shell interactors were set as white nodes. Edges indicate known molecular action of a protein node regarding another protein node. ACE2 is directly connected to components of the kinin-kallikrein (bradykinin), renin-angiotensin and coagulation systems.

Angiotensin converting enzyme 2 (ACE2) is the receptor for SARS-CoV-2 (*8*). Binding occurs through viral spike protein (*9*) and depends on the serine protease TMPRSS2 for priming (*8*). Physiologically, ACE2 is involved in the control of three independent but highly integrated systems, the kinin-kallikrein (KKS), renin-angiotensin (RAS), and coagulation (CS) systems (Fig. 1B). Bradykinin is a potent inflammatory substance produced from high-molecular weight kininogen (HMWK) in a reaction catalyzed by the serine protease kallikrein (*10*). Bradykinin can directly deliver its vasoactive and inflammatory actions through bradykinin receptor 2 or be further processed by carboxypeptidase N to form DR9-bradykinin that activates bradykinin receptor 1 to deliver inflammatory and pain signals (*11*). ACE2 degrades DR9-bradykinin into inactive peptides and, together with angiotensin converting enzyme (ACE), which inactivates bradykinin, shuts down the KKS (*12*). Angiotensin II (Ang II) is a pleotropic hormone involved in the regulation of blood pressure, blood volume, cardiac function, and electrolyte balance (*13, 14*). It is produced from angiotensin I (Ang I) through the catalytic action of ACE, whereas ACE2 catalyzes its degradation into the inactive peptide angiotensin 1-7, thereby inactivating the RAS (*15*). The interaction of ACE2 with the CS is indirect and occurs via two mechanisms: 1) catalyzing the production of angiotensin 1-9, which reduces plasminogen activator and increases PAI-1, thus inhibiting fibrinolysis (*16*) and 2) modulating the activity of kallikrein, which in turn catalyzes the conversion of plasminogen into plasmin (*17*).

Upon SARS-CoV-2 binding, ACE2 is internalized to endosomes, leading to a subcellular location shift that could alter its capacity to physiologically regulate the KKS, RAS and CS (*18-22*). This could simultaneously impact the highly lethal COVID- 19 triad: lung inflammation, cardiovascular failure, and coagulopathy. Despite the fact that ACE2 is expressed in several organs and tissues, both clinical and experimental evidence shows that SARS-CoV-2 promotes most of its pathological actions by initially infecting cells of the upper respiratory tract and, subsequently, alveolar cells in the lung (*23-25*). It is currently unknown if lung cells expressing ACE2 are equipped with proteins that belong to the KKS, RAS and CS, which could potentially explain a cell-autonomous system that is disturbed by SARS-CoV-2 infection, leading to abnormal regulation of all three systems. Here, we evaluated the transcripts of 129,079 human lung cells previously submitted to single-cell RNA sequencing. We show that transcripts encoding for key elements of all three systems are highly co-expressed with ACE2 in alveolar cells.

## Results

### ACE2 interacts with proteins belonging to the kinin-kallikrein, renin-angiotensin, and coagulation systems

A protein-protein interaction network (Fig. 1C) revealed that ACE2 interacts closely with proteins that belong to the KKS; kininogen (KNG1), the substrate for bradykinin synthesis, and kallikrein (KLKB1), the enzyme that catalyzes this conversion. ACE2 also interacts with proteins of the RAS; ACE, that converts Ang I into Ang II, renin (REN), the enzyme that converts angiotensinogen (AGT) into Ang I, AGT itself and angiotensin receptor 1 (AGTR1). The interface of ACE2 with CS occurs mostly though KLKB1 that controls fibrinolysis; in addition, Factor II (F2, thrombin), was identified in the interactome.

### Integrated analysis of 129,079 human single lung cells leads to the identification of cell types expressing ACE2

To investigate the lung cell types that are potentially targeted by SARS-Cov-2 due to ACE2 expression, we leveraged public single-cell RNA sequencing (scRNAseq) from three previously published datasets and a pre-print report (*26-28*) (https://doi.org/10.1101/742320). Despite active investigation of the cellular landscape of the human lung, the field lacks an integrated atlas in which a consensus can be established between various datasets. We addressed this issue by individually filtering and integrating each study control sample into a batch-corrected study-wise reference with Seurat v3 anchor-based integration (Fig. 2A, Suppl. Fig. 1, Suppl. Fig. 2A-B, Suppl. Table 1). These batch-corrected data were used for integration of peer-reviewed studies. Preprint data was used for cell-type annotation by by anchor transferring learning (https://doi.org/10.1101/742320). Corrected data were dimensionality reduced with diffusion-based Manifold Approximation and Projection (dbMAP) (http://dx.doi.org/10.2139/ssrn.3582067), a dimensionality reduction method tailored for the analysis of large-scale single-cell data and clustered by its diffusion graph structure (Fig. 2B). In this embedding, each point represents a single-cell, and the position in the embedding represents its relative transcriptional identity when compared with other cells. Grouping cells by study of origin shows that this approach is successful in removing experimental noise from data (Suppl. Fig. 2B). Cells were clustered by applying the Louvain algorithm to the graph of each cell diffusion structure scoring. After removal of singletons and doublets, 47 cell clusters were identified, comprising eight major groups: alveolar cells, endothelial and lymphatic cells, monocytes/macrophages, T cells, B cells, fibroblasts, smooth muscle cells, and mast cells (Fig. 2B). These clusters were annotated based on learned annotations published elsewhere (https://doi.org/10.1101/742320) (Suppl. Fig. 2C) and on cluster marker gene expression (Suppl. Fig. 3). The clusters effectively correspond to biologically comprehensive cellular states that can be explored as a resource to COVID-19 studies. This potential is leveraged by dbMAP for the identification of rare cell types and cellular-state transitions in comparison against UMAP (Suppl. Fig. 2D). An illustrative example is shown by B-cell differentiation into plasma cells. In the dbMAP embedding, B and plasma cells form discrete populations connected directly by intermediate cells (Fig. 2B), whereas UMAP represents these clusters as completely separated populations (Suppl. Fig. 2D). We briefly explored cluster marker genes (Suppl. Figs. 4 and 5), albeit the exploration of this atlas exceeds the scope of this manuscript. An interactive browser with our data is available at https://humanlung.iqm.unicamp.br, allowing non-bioinformaticians to produce publication-level plots and tables in a community effort to further explore the human lung cellular landscape. We next investigated the expression of ACE2 We then investigated the expression of ACE2 and show that it is restricted to alveolar cells and fibroblasts, being practically absent from other cell clusters (Fig. 1C).

**Figure 2.**
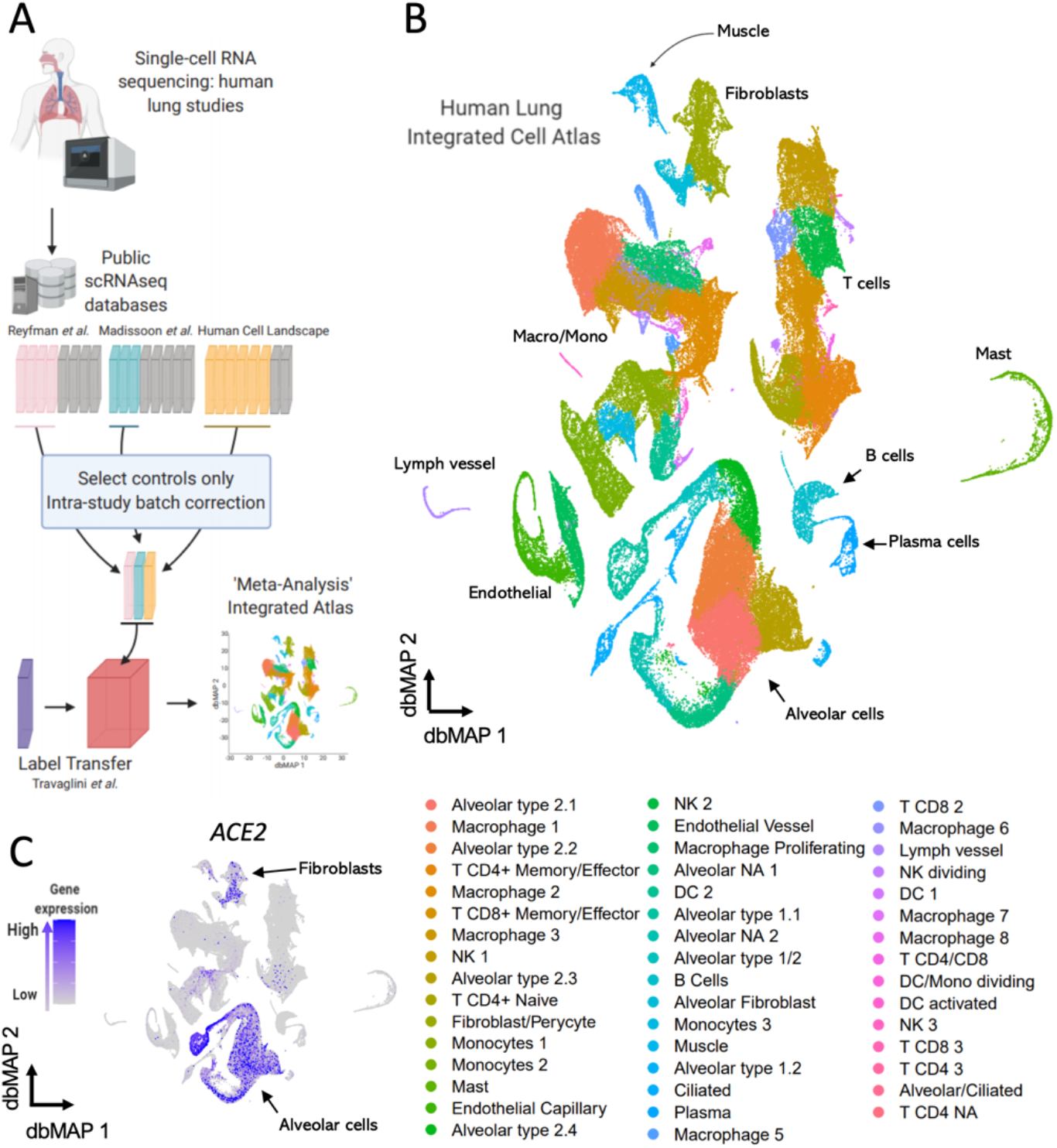
Generation of an Integrated Lung Cell Atlas through integration of public data reveal fibroblasts and alveolar cells expressing ACE2 in the human lung. **a**. Public data from single-cell RNA sequencing (scRNAseq) studies on the human lung was retrieved. Reyfman *et al*., Madissoon *et al*. and Human Cell Landscape control datasets were individually analyzed and batch-corrected with Seurat v3 anchor-based integration after filtering and normalization. Batch-corrected data representing each study was used to assemble an integrated dataset. This integrated dataset was annotated with the assistance of label transferred from Travaglini *et al*. data. **b**. The Human Lung Integrated Cell Atlas represented in two-dimensional diffusion-based Manifold Approximation Embedding (dbMAP). dbMAP organizes the visualization to preserve as much of the original data structure as possible in a comprehensive way. In this representation, each cell is a point mapped to an embedding so that its ***x, y*** coordinates represent its relative transcriptional identity, i.e. its phenotypic signal. Cells are colored by their assigned cluster. **c**. Visualization of ACE2 expression on the human lung dbMAP embedding. ACE2 is consistently expressed in alveolar cells and scattered in fibroblasts, being practically absent from other cell types. NK = Natural Killer cell; DC = Dendritic Cell; NA = Not Assigned.

### TMPRSS2 is co-expressed with ACE2 in alveolar cells

The serine protease TMPRSS2 is required for SARS-CoV-2 spike protein priming and subsequent binding to ACE2 (*8*) (Fig. 3A). Following receptor binding, SARS-CoV-2 is internalized through endocytosis in a process that depends on the activity of phosphatidylinositol 3- phosphate 5-kinase (PIKFYVE) and its downstream effector, two-pore channel subtype 2 (TPCN2) (*18*) (Fig. 3A). Moreover, the inhibition of cathepsin L (CTSL) dramatically reduces virus entry, suggesting a role for this lysosome protein in the process (*18*) (Fig. 3A). Out of the four proteins currently described as players in SARS-CoV-2 cell entry, only TMPRSS2 gene expression selectively overlapped with ACE2 expression (Fig. 3B, 3C, and 3G). TMPRSS2 transcripts are expressed in virtually all alveolar cell clusters in addition to ciliated and alveolar/ciliated clusters. As expected for a phosphatidylinositol 3-phosphate kinase, PIKFYVE is ubiquitously expressed throughout all lung-cell clusters, with some predominance in clusters of alveolar type 2 cells, B-lymphocytes, and mast cells (Fig. 3D and 3G). Likewise, TPCN2 is ubiquitously expressed in lung-cell clusters, albeit in fewer cells than PIKFYVE and some predominance in T-lymphocytes and NK cells (Fig. 3E and 3G). CTSL is also expressed in virtually all lung-cell clusters. However, here there is a large predominance in macrophage clusters (Fig. 3F and 3G).

**Figure 3.**
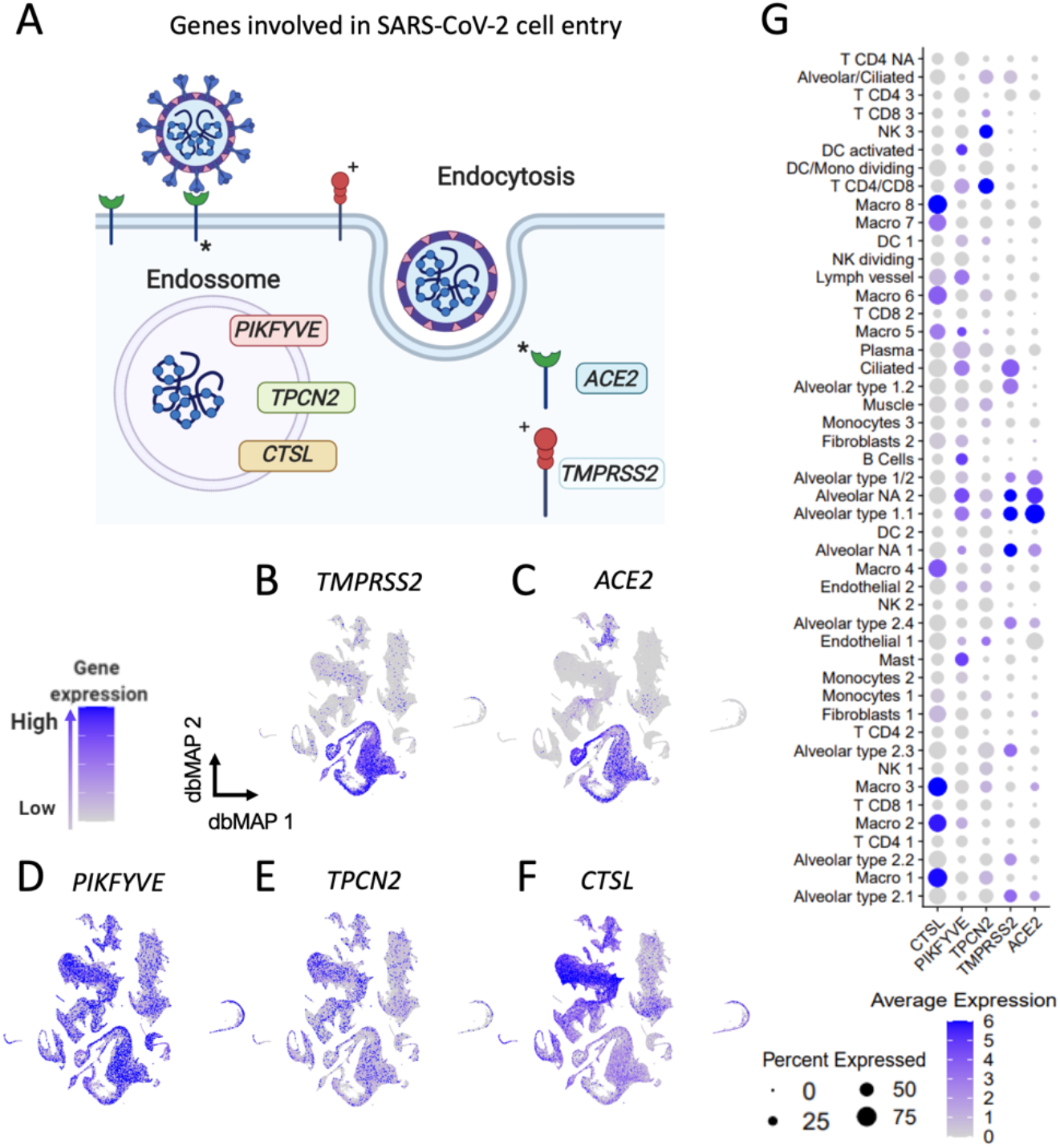
Lung expression of genes involved in SARS-Cov-2 cell entry is selective to alveolar cells. **a**. Schematic representing SARS-Cov-2 cell entry mechanism. TMPRSS2 cleaves SARS-Cov-2 *spike* protein, allowing it to bind to ACE2, its functional receptor. It is known that other genes such as PIKFYVE, TPCN2 and CTSL also play a role in SARS-Cov-2 endocytosis and cellular contamination **b-f**. Visualization of gene expression in dbMAP embeddings of the lung atlas. **b**. TMPRSS2 expression is restricted to alveolar cells. **c**. ACE2 expression is selective to alveolar cells and fibroblasts. **d-f**. PIKFYVE, TPCN2 and CTSL are expressed consistently throughout the majority of lung cell types. CTSL expression in macrophages **g**. Dot plot visualization of expression of genes involved in SARS-Cov-2 cell entry in human lung cell types. ACE2 expression is selective to alveolar clusters, similarly to TMPRSS2, but with the particularity of being lowly expressed in a large portion of endothelial cells. PIKFYVE, CTSL and TPCN2 are expressed by all clusters, but CTSL expression is much higher in macrophages, while TPCN2 expression is higher in NK and T CD4/CD8 clusters.

### Kininogen is co-expressed with ACE2 in alveolar cells

Kininogen (KNG1), the precursor for bradykinin synthesis, is expressed in a large number of cells that also express ACE2 (Fig. 4A1, 4A6, and 4B). KNG1 expression is restricted to alveolar type 1.1, type 1/2, and type 2.4, in addition to alveolar 2.2. and 2.3 clusters. Kallikrein (KLKB1) is a serine protease that catalyzes the conversion of kininogen into bradykinin. It is expressed predominantly in endothelial and lymph vessels cells (Fig. 4A2 and 4B). KLKB1 is also expressed in scattered alveolar cells and in some T-lymphocytes and macrophages (Fig. 4A2). Bradykinin acts through the type 2 cognate receptor (BDRK2), which is expressed predominantly in endothelial vessel cells and in some alveolar cells, particularly types 1.2 and 2.3 (Fig. 4A3 and 4B). BDRK2 is also expressed in a considerable number of fibroblasts and plasma/B-lymphocytes (Fig. 4A3 and 4B). Bradykinin can either be converted into inactive kinins by the catalytic action of ACE or be converted into the active DR9-bradykinin. ACE is expressed predominantly in macrophages, monocytes, and endothelial cells (Fig. 4A4 and 4B). It is also expressed in scattered alveolar cells, T-lymphocytes, and fibroblasts (Fig. 4A4 and 4B). DR9-bradykinin acts through bradykinin receptor 1 (BDRK1), which is predominantly expressed in fibroblasts, endothelial cells, and alveolar type 2.1, 2.2, and 2.3 cells (Fig. 4A5 and 4B).

**Figure 4.**
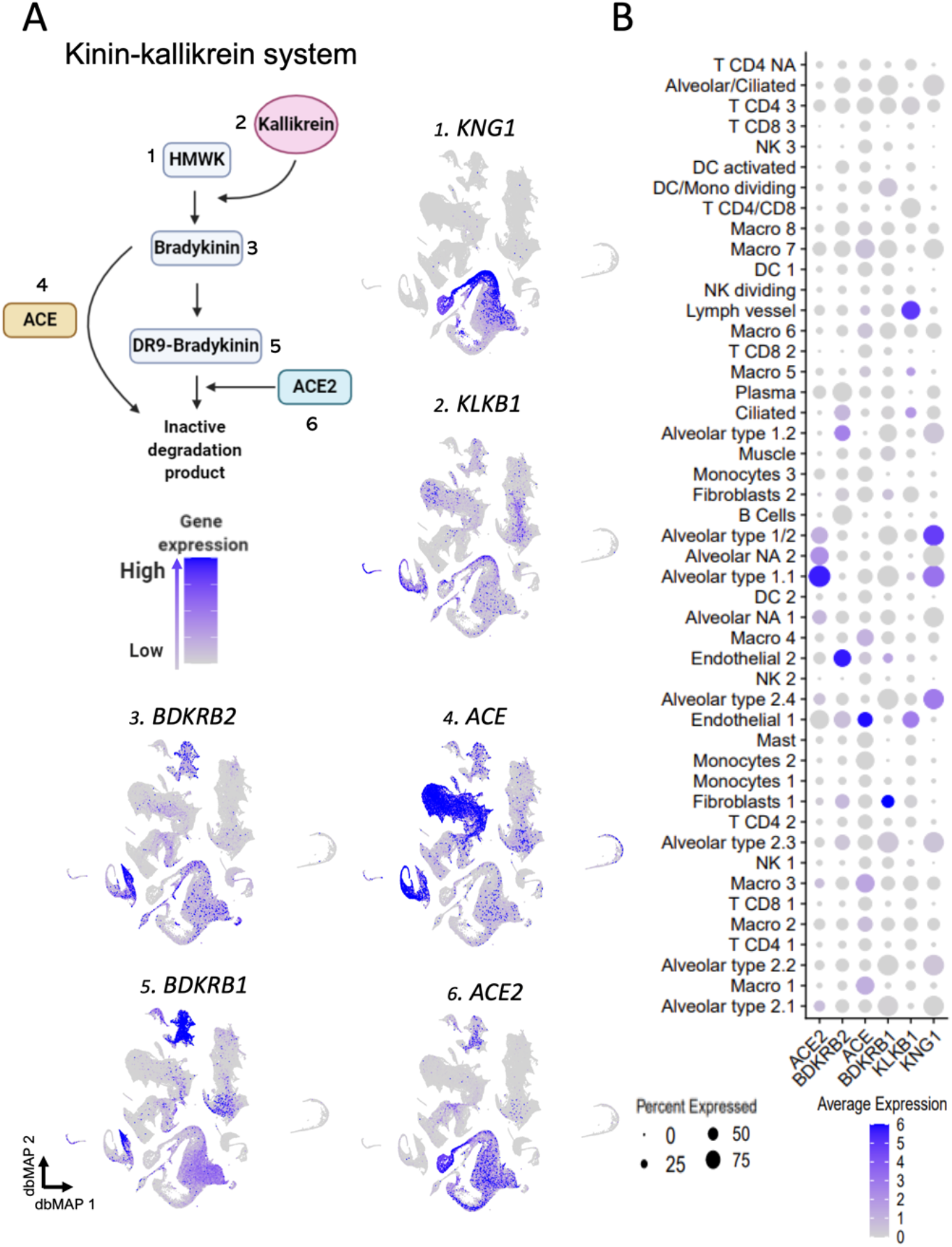
Gene expression of components of the kinin-kallikrein system point to alveolar cells role. **a**. Schematic representing the main players and steps of the bradykinin system, also known as the kinin-kallikrein system. High molecular weight kininogen (HMWK) conversion into bradykinin is catalyzed by kallikrein. Bradykinin binds to the bradykinin receptor 2 (BDKRB2), but it can be converted into DR9-bradykinin, which activates the bradykinin receptor 1 (BDKRB1) until it is degraded by ACE2 into inactive degradation products. Bradykinin can also be directly converted into inactive degradation products by ACE. **1-6**. Visualization of gene expression in dbMAP embeddings of the lung atlas. **1**. KNG1 is selectively expressed in alveolar cells. **2**. KLKB1 is expressed in endothelial and lymph vessel cells, and sparsely expressed in alveolar cells. **3**. BDKRB2 expression is selective to endothelial cells. **4**. ACE expression is selective to endothelial cells and macrophages. **5**. BDKRB1 expression is selective to endothelial cells and fibroblasts, but also detected in low levels in alveolar cells. **6**. ACE2 expression is restricted to alveolar cells. **b**. Dot plot visualization of expression of genes involved in the bradykinin system in the lung atlas cell types. ACE2 and KNG1 gene expression is selective to alveolar clusters, despite ACE2 being lowly expressed in a large portion of endothelial cells and KNG1 also being lowly expressed in a large portion of clusters of macrophages. KLKB1 is highly expressed in endothelial and lymph vessel cells, but also lowly expressed in T CD4 clusters. BDKRB2 is highly expressed in endothelial and alveolar type 1.1 cells, and also less expressed by a large portion of B/plasma cells, and of macrophages clusters 7 and 8. ACE is selectively expressed in endothelial cells and in macrophages. BDKRB1 expression is selective to fibroblasts, although various clusters express it in low levels. ACE2 expression is restricted to alveolar cells.

### Renin and angiotensin receptor 1 are co-expressed with ACE2 in alveolar cells

Angiotensinogen (AGT) is the precursor for Ang I; we show that most cells expressing AGT are fibroblasts and smooth muscle cells; however, a considerable number of type 2.4, 2.3, and 2.1 alveolar cells that express ACE2 also express AGT (Fig. 5A1, 5A2, and 5B). Renin is the enzyme that converts AGT into Ang I; here, we show that in the lung it is predominantly expressed in alveolar cells and largely co-expressed with ACE2, particularly in alveolar cell types 1.2, 2.2, 2.3, 2.4, and NA 2 (Fig. 5A1, 5A3, and 5B). Some macrophages and mast cells also express renin (Fig. 5A3 and 5B), which is virtually absent in the remaining clusters. Ang I is converted into active Ang II by ACE (Fig. 5A4 and 5B), which is largely expressed in macrophages and endothelial cells. Ang II exerts most of its cardiovascular effects by acting through angiotensin II receptor 1 (AGTR1). We show that most cells expressing AGTR1 are fibroblasts and muscle cells (Fig. 5A5 and 5B); in addition, a considerable number of alveolar type 1.1 and NA 2 cells that express ACE2 also express AGTR1 (Fig. 5A1, 5A5, and 5B).

**Figure 5.**
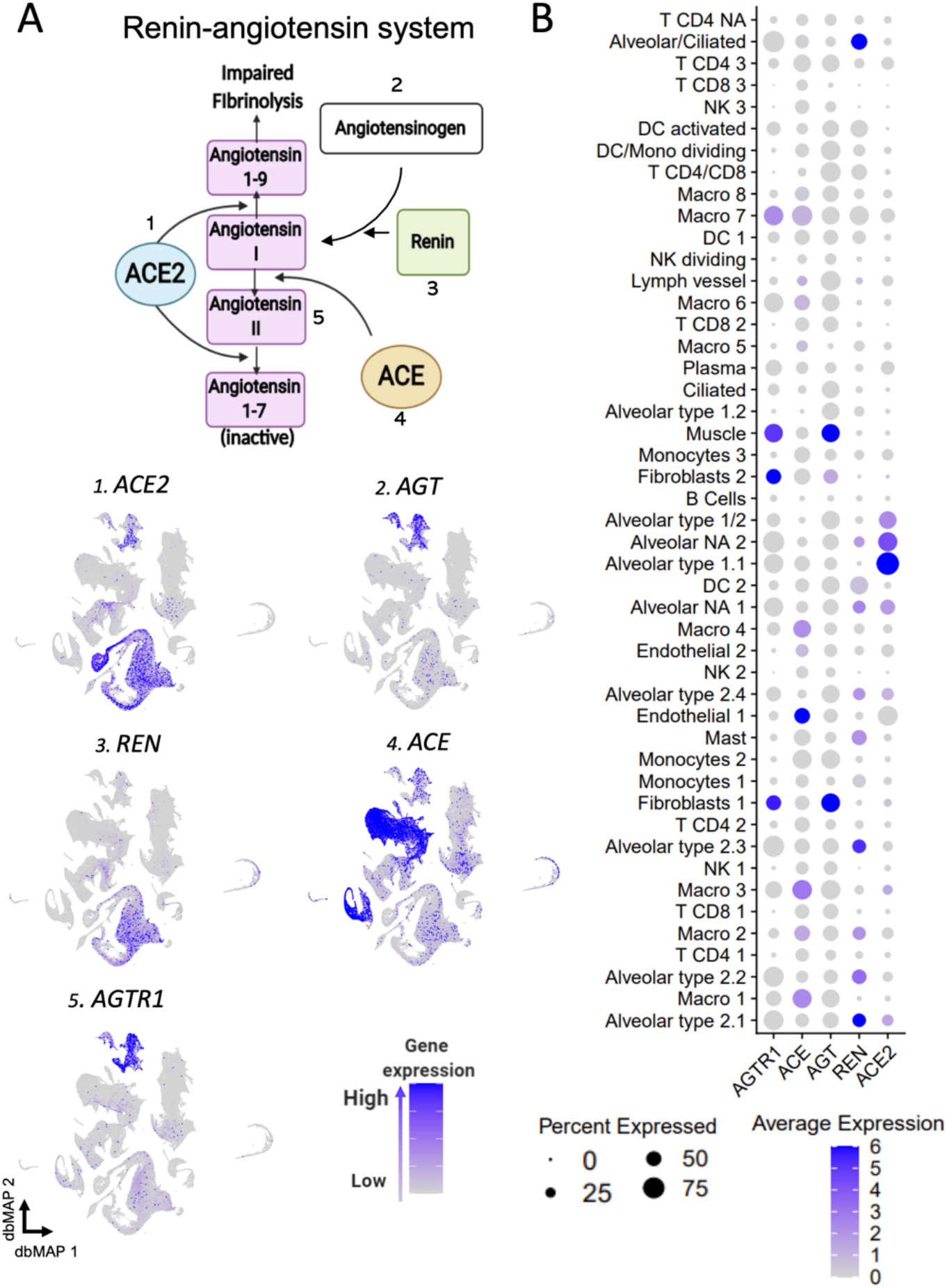
Gene expression of components of the renin-angiotensin system point towards alveolar cells role. **a**. Schematic representing the main players and steps of the angiotensin system, also known as the renin-angiotensin system (RAS). ACE2 catalyzes the conversion of angiotensin I to angiotensin 1-9, and of angiotensin II into inactive angiotensin 1-7. Renin catalyzes the conversion of angiotensinogen into angiotensin I. ACE catalyzes the conversion of angiotensin I into angiotensin II, which binds to its receptor AGTR1. **1-5**. Visualization of gene expression in dbMAP embeddings of the lung atlas. **1**. ACE2 expression is restricted to alveolar cells. **2**. AGT is expressed in fibroblasts and smooth muscle cells. **3**. REN expression is restricted to alveolar cells. **4**. ACE expression is selective to endothelial cells and macrophages. **5**. AGTR1 expression is selective to fibroblasts and smooth muscle cells. **b**. Dot plot visualization of the expression of genes involved in the angiotensin system in the lung atlas cell types. ACE2 expression is restricted to alveolar cell clusters, similarly to REN, which is virtually absent from remaining clusters. AGT is highly expressed in fibroblasts and muscle, albeit being lowly expressed in a large portion of cells from other clusters. ACE is preferentially expressed in endothelial cells and macrophage clusters. AGTR1 is highly expressed in smooth muscle cells and fibroblasts, and lowly expressed in macrophages and alveolar cells.

### Fibrinogen gamma is co-expressed with ACE2 in alveolar cells

In addition to its action on the KKS, kallikrein (KLKB1) acts in the CS by converting plasminogen into the fibrinolytic substance plasmin; KLKB1 is expressed predominantly in endothelial and lymph vessel cells and in alveolar type 1.1, type 1/2, and type 2.4 cells (Fig. 6A1 and 6B). Another catalyst of plasmin production is tissue plasminogen activator (PLAT), which is inhibited by PAI-1 (SERPINE1). SERPINE1 is expressed predominantly by endothelial cells and secondarily by fibroblasts and muscle cells (Fig. 6A2 and 6B), whereas PLAT is expressed in endothelial and lymph vessel cells (Fig. 6A3 and 6B). Fibrinogen gamma chain (FGG) is the substrate for production of fibrin, the main component of clot formation; it is expressed in virtually all alveolar cell types except for types 1.1 and 1.2, thus largely coinciding with the expression of ACE2 (Fig. 6A4, 6A5, and 6B).

**Figure 6.**
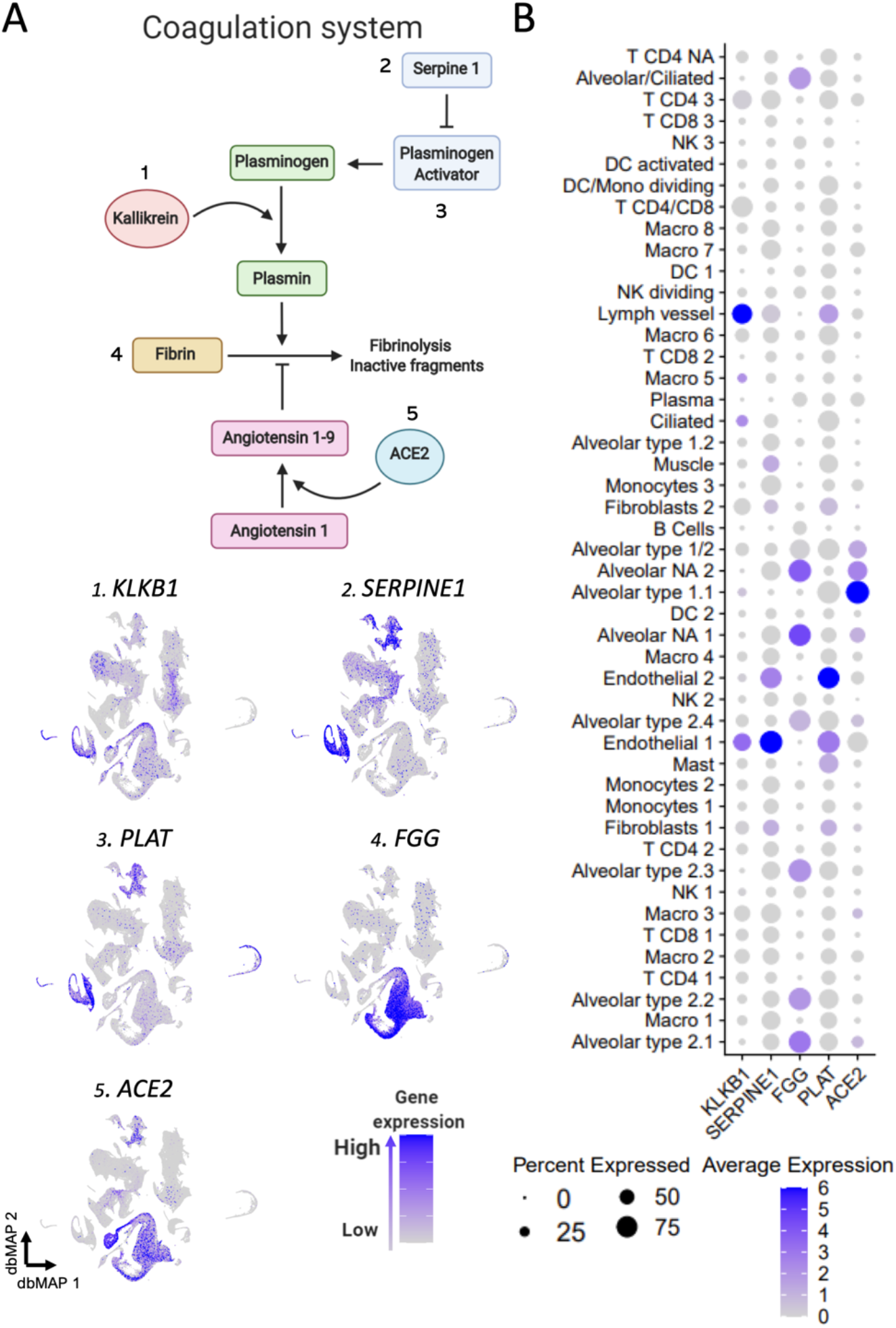
Gene expression of components of the coagulation system. **a**. Schematic representing the final step of the coagulation system. Plasma kallikrein (KLKB1) catalyzes the conversion of plasminogen into plasmin. Plasminogen is activated by tissue plasminogen activator (PLAT), which is regulated by serpine 1 (SERPINE1). Plasminogen conversion into plasmin promote fibrinolysis by degradation of fibrin networks generated from fibrinogen (FGG) into inactive fragments. ACE2 plays a role in this process by catalyzing the conversion of angiotensin I to angiotensin 1-9, which inhibits fibrinolysis. **1-5**. Visualization of gene expression in dbMAP embeddings of the lung atlas. **1**. KLKB1 is expressed in endothelial cells and sparsely in alveolar cells. **2**. SERPINE1 is highly expressed in fibroblasts and in endothelial and smooth muscle cells, and present in lower levels in macrophages. **3**. PLAT is mostly expressed in endothelial cells and fibroblasts. **4**. FGG expression is restricted to alveolar cells. **5**. ACE2 expression is restricted to alveolar cell clusters. **b**. Dot plot visualization of the expression of genes involved in the coagulation system in the lung atlas cell types. ACE2 expression is restricted to alveolar cell clusters. PLAT expression is higher in endothelial cells, lymph vessel cells and mast cells. FGG expression is restricted to alveolar cell clusters, being higher in alveolar type 2.1 and type 2.2, alveolar/ciliated and alveolar NA clusters, a poorly characterized cellular population.

## Discussion

Here, we present the largest single-cell analysis of the human lung cellular transcriptional landscape to date. This was achieved by assembling datasets provided by previous independent studies into an integrated human lung cell atlas containing 129,079 cells (*26-28*). We further leveraged the power of this data by analyzing it with dbMAP, a novel dimensional reduction method that, together with the integrated dataset, are publicly available at (https://github.com/davisidarta/humanlung and https://github.com/davisidarta/dbMAP). An user-friendly interactive web-based platform is also available at https://humanlung.iqm.unicamp.br, in a powerful data exploration environment that holds potential to accelerate lung research. This data can also be used by future studies performing scRNAseq and data analysis of the human lung, so that others may have the option to add their study into this integrated atlas. By using this integrative. Using this approach, we confirmed previous data that identified alveolar cells as those expressing highest levels of the SARS-CoV-2 receptor, ACE2, in the lung (*23*). In addition, we identified cell types that express transcripts encoding for proteins potentially involved in SARS-CoV-2 cell entry. We demonstrated that expression of TMPRSS2, a serine protease that primes SARS-CoV-2 spike protein, largely overlaps with ACE2 expression in alveolar cells (*23*), reinforcing the hypothesis of TMPRSS2 as a promising pharmacological target of COVID-19 (*29*). Moreover, we showed that transcripts encoding for PIKFYVE, TPCN2, and CTSL (*18*) are ubiquitously expressed in the lung and, even though they are expressed in alveolar cells with some degree of overlap with ACE2, their potential as therapeutic targets could be challenged due to lack of cell specificity.

Next, we determined the identities of lung cells expressing key components of the KKS, RAS, and CS pathways. Currently, there is neither experimental nor clinical evidence suggesting that SARS-CoV-2 infection could lead to bradykinin-dependent lung inflammation and edema. However, in a rodent model of lung LPS-induced inflammation, the inhibition of ACE2 resulted in a significant increase lung inflammatory edema, and the simultaneous inhibition of bradykinin in this model dramatically reduced inflammatory damage (*12*). In addition, in humans, severe lung edema can develop in a rare genetic disorder, hereditary angioedema, that results from bradykinin accumulation due to mutations of the C1 esterase/kallikrein inhibitor (*30*). Clinical studies have shown that inhibitors of bradykinin can efficiently treat lung edema in these circumstances (*31*). Furthermore, the use of ACE inhibitors provides yet another line of clinical evidence for disturbed lung accumulation of bradykinin that can, on rare occasions, lead to severe outcomes (*32, 33*). Here, we showed that kininogen, the precursor of bradykinin, presents considerable cellular expression overlap with ACE2 and that transcripts encoding for other components of the KKS are also expressed in lesser amounts in ACE2-expressing cells.

In contrast to KKS, abnormalities in the RAS have been widely reported in COVID-19 patients (*34-36*). Obesity, hypertension, diabetes, and coronary insufficiency represent the greatest risk factors for severe COVID-19 (*5, 6*), and in all these diseases, there is an increased risk for abnormal regulation of the RAS (*13, 37, 38*). In one of the largest series reporting COVID-19 patients published so far, the use of ACE inhibitors before infection was shown to reduce mortality by 33%, representing the most effective independent factor, among those analyzed, that could protect patients from a lethal outcome (*6*). Here we showed that angiotensinogen, the precursor of Ang 1, and particularly renin, the enzyme catalyzing this conversion, are expressed in alveolar cells and that their expression overlaps that of ACE2. In addition, other key components of the RAS, ACE, and angiotensin receptor 1, are also co-expressed with ACE2 in a considerable number of cells.

COVID-19-associated coagulopathy can lead to a fulminant activation of coagulation, resulting in widespread thrombosis. Venous thromboembolism (VTE) is one of the leading causes of severe complications in COVID-19 patients (*3*, *39*). It was diagnosed in 20% of patients admitted to an intensive care unit (ICU) and its cumulative incidence increased progressively to 42% as patients remained in severe condition in the ICU (*39*). Developing VTE during the progression of severe COVID-19 increases the risk of death by 140% (*39*). Moreover, D-dimer, a degradation product of fibrin, has been identified as an important predictive marker of severe COVID-19 and was directly correlated with poor prognosis (*40, 41*). Because of the association between disturbed coagulation and severe disease progression, the use of anticoagulant treatment has been proposed as potentially beneficial in COVID-19 (*22, 42*). In one therapeutic clinical trial, the use of low-molecular-weight heparin resulted in reduced mortality in patients with high levels of D-dimer (*43*); however, in another study, preventive anticoagulation was associated with increased VTE (*44*). These results suggest that prophylactic and therapeutic anticoagulation approaches have distinct outcomes in COVID-19; however, further studies are awaited in order to direct the establishment of optimal measures that could lessen the severity of COVID-19 coagulation abnormalities. Here, we showed that particularly FGG and KLKB1 are expressed in alveolar cells that also express ACE2. Both FGG and KLKB1 play central roles in the control of the CS. FGG deficiency leads to mild or even severe bleeding (*45*), whereas KLKB1 deficiency is associated with an increased risk of thrombosis (*46*).

In conclusion, alveolar cells expressing ACE2, which are primary cellular targets for SARS-CoV-2 in the lung, also express transcripts encoding proteins that play pivotal roles in the regulation of the KKS, RAS, and CS. As all these systems are potentially affected during the progression of severe COVID-19, we propose that abnormal function of ACE2 as a result of SARS-CoV-2 infection could directly and cell-autonomously precipitate the development of acute lung inflammation, cardiovascular failure and thromboembolism, which are the hallmarks of severe COVID-19 (*46*).

## Methods

### Computational Environment

In silico analyses were performed on two different machines. Locally, a ThinkPad P52 Workstation with 128GB RAM and a six-core Intel Xeon processor was used for data exploration. A high-performance computing cluster (HPCC) was used for data integration and computation of results. Analysis was performed in R version 3.6.2. A docker environment with a pre-installed RStudio image and loaded with all required packages is available at https://github.com/davisidarta/lungcovid.

### Protein-Protein Interaction Networks

Protein functional interaction networks were performed with STRING v11 (*47*), a software toolkit which performs datamining on a large number of databases and on individually published high-throughput datasets. The default functional interaction network was queried for ACE2 in the *Homo sapiens* organism and was visualized by known molecular action. We subsequently added a second shell of interactors in order to explore deeper interactions through autonomous datamining. A permalink webpage of ACE2 Protein-Protein Interaction network is accessible through https://version-11-0.string-db.org/cgi/network.pl?networkId=KwL0Kho7pBZf.

### Single-cell RNA sequencing data acquisition, filtering, and processing

Control samples (healthy donors) digital expression matrices were downloaded from Gene Expression Omnibus for data published elsewhere (*26*) (GSE122960) and for Human Cell Landscape (HCL) data (*27*) (GSE132355). For other datasets (*28*), raw digital expression matrices and associated metadata were downloaded from the Human Cell Atlas web portal (https://data.humancellatlas.org/explore/proiects/c4077b3c-5c98-4d26-a614-246d12c2e5d7). In addition, preprint data from Travaglini and coworkers (https://doi.org/10.1101/742320) was obtained from synapse (Human Lung Cell Atlas – https://www.synapse.org/#!Synapse:syn21041850/wiki/600865). Cells were filtered by total number of reads (*nreads*), by number of detected genes (*ngenes*), and by mitochondrial percentage (*mito.pc*). Data filtering thresholds were defined by inflection points in the logarithmic curves of each cell’s number of reads (*nreads*) and number of detected genes (*ngenes*). Filtering was performed on each individual sample prior to integration, with the exception of the study by Madissoon *et al*., in which each sample corresponds to a different point in a time course of frozen storage of cells prior to processing (0 h-72 h), rendering extremely low batch effects. Filtering was performed in each individual sample prior to integration, with the exception of Madissoon *et al*. study, in which each sample corresponds to a different point in a time-course of frozen storage of cells prior to processing (0h – 72h), rendering extremely low batch effects. Supplementary Table 1 summarizes the thresholds used by each sample in each individual study, and Supplementary Figure 1 summarizes quality-control metrics for each dataset.

Each individual sample was processed in Seurat v3.1.5 (*48*) using the default Seurat workflow. For each individual sample, cell counts were log-normalized by a size factor of 10,000 RNA counts, and feature selection was performed by selecting the 5,000 genes with the highest dispersion. Data were then scaled (z-core transformation to standardize expression values for each gene across all cells). For sample batch-correction within each study, we performed the combination of the gene expression of each sample and then selected 2,000 features to be included in the integrated analysis prioritizing features that were identified as highly variable in multiple samples.

Unsupervised identification of anchor correspondences between the Canonical Correlation Analysis (CCA) space of each sample normalized data was performed with the *FindIntegrationAnchors* function in Seurat v3 with default parameters. These anchors were scored and weighted, being subsequently merged into integrated assays containing the balanced expression of the union of each sample’s expressed genes, resulting in three study-wise balanced datasets.

### Data integration and label transfer

For integration of data from different studies, we performed the union of the gene expression of available human lung peer-reviewed studies (*26-28*). Prior to anchoring, 3,000 features were selected by ranking top features identified as highly variable in multiple studies. We proceeded similarly to the individual study level, where anchors were built between multiple samples, and searched for anchor correspondences between each study’s own manifold by using Seurat v3 Integration. As a first step, dimensional reduction with CCA is performed. CCA effectively captures gene modules that are correlated and shared in pair-wise dataset comparisons, representing signals that define a shared biological state. Afterwards, mutual nearest neighbors (MNN – pairs of cells, each from one dataset) that share a corresponding state are weighted in an ‘anchor’, which is scored so as to exclude spurious connections between unrelated cell types. The integration of these anchors into a single manifold was performed with the *IntegrateData* function, and the top 20,000 genes with higher dispersion were included in the final result. The data of Travaglini *et al*. were not included in the integrated atlas due to its lack of peer review. However, we did use these data annotations for label transfer due to its high-quality cell-type annotation. This was performed by employing the FindTransferAnchor and TransferData functions, in which a classification matrix is transposed by multiplication by a weighting matrix to return prediction scores for each class for every cell in the atlas. Those labels were further used as guidance in the process of cell-type annotation, as well as the expression of cell-type marker genes.

### Dimensionality reduction with diffusion-based Manifold Approximation and Projection (dbMAP)

Visualizing single-cell data is a challenging task in which comprehensive two- or three-dimensional embedding needs to be generated from an exceptionally large number of samples and observations. Previous approaches mainly relied on performing prior dimensionality reduction with PCA to denoise data and make it computationally easier to compute non-linear dimensional reduction methods such as t-stochastic neighborhood embedding (t-SNE), UMAP, and potential of heat diffusion for affinity-based trajectory embedding (PHATE). We have recently proposed diffusion-based Manifold Approximation and Projection (dbMAP), which excels at identifying rare cell populations and describing lineage dynamics as trajectories progress, branch, and cycle (http://dx.doi.org/10.2139/ssrn.3582067). Briefly, dbMAP encodes and denoises data by dissecting its diffusion structure, in which cell-cell similarity information is adaptive regarding each cell’s individual neighborhood density. This information is then propagated through a series of random walks, which are scaled to normalize the relative progression of diffusion during each walk. This approach effectively adapts diffusion maps to approximate the Laplace-Beltrami operator, which represents data intrinsic structure. The resulting eigenvectors are diffusion components which can be used for downstream clustering, layout visualization and pseudotime estimation, being a potential substitution for PCA in single-cell processing. These diffusion components address UMAP’s assumptions on its input, thus optimizing its potential for visualization, a feature we explored with dbMAP. dbMAP is computationally scalable and robust to variations in parameters, uniquely allows the visualization of cyclic trajectories in which stem cells actively traverse the cell cycle and is well-suited for the analysis of organ- and organism-level data. dbMAP takes four main parameters. During diffusion, a number **N** of structure components are computed accounting for each cell **K** nearest neighbors. After automatic scaling and selection of relevant components by eigengap analysis, a UMAP layout is generated with **M** as the effective minimum distance between embedded points and **S** as the effective scale of embedded points. Importantly, visualization parametrization can be fine-tuned by the user for its specific dataset due to the fast UMAP layout computation of the structure components, although results overall are robust to small changes in these parameters. Parameters used for dbMAP embedding for individual studies and the integrated atlas, as well as those used for UMAP embedding of the atlas, are listed in Supplementary Table 3.

### Clustering

Clustering was performed by generating a k-nearest-neighbors graph from the structure components learned in dbMAP first step. For this, we applied the FindNeighbors function in Seurat with default parameters on the structure components. Clustering was then performed by a shared nearest neighbor (SNN) modularity-based clustering algorithm by using the FindClusters function in Seurat with default parameters.

### Identification of marker genes

Cluster marker genes were found with Seurat’s function FindAllMarkers, which finds differentially expressed genes between a cluster and all remaining cells with a Wilxocon rank sum test. Marker genes were then ranked by their log of fold change of expression in particular cell types. The two marker genes with the highest log of fold change for each cluster were chosen for the Dotplot and heatmap visualizations. A comprehensive table of all marker genes is provided as Supplementary Data.

### Cell-type annotation

Clusters were annotated to cell types corresponding the metadata of Madissoon *et al*. and the transferred labels from Travaglini *et al*. and accordingly to canonical marker gene expression.

### Doublet identification and removal

We identified five clusters that presented without specific marker genes and in spurious trajectories as doublets and removed them. Three clusters scattered the white space between alveolar cells, monocytes/macrophages, and lymphocytes in pairwise trajectories between these three cell types, effectively corresponding to doublets. The two remaining doublet clusters were scattered between mast cells and monocytes. We provide code for the identification and removal of these doublet clusters.

### Visualization of gene expression

Visualization of gene expression was performed with dbMAP plots of gene expression and with dot plots. The Seurat functions FeaturePlot and DotPlot were applied to the integrated Seurat object with particular coloring thresholds for each gene so as to visualize expression throughout the color scale (Suppl. Table 4).

## Data Availability

All used data is publicly available. The integrated atlas is publicly available at https://humanlung.iqm.unicamp.br for exploration as a downsampled Cerebro object. The full integrated data and code used in the analysis are available at https://github.com/davisidarta/humanlung.

https://humanlung.iqm.unicamp.br

## Code Availability

All code used for analysis of single-cell RNA sequencing data is available at https://github.com/davisidarta/humanlung. dbMAP is available at https://github.com/davisidarta/dbMAP as a python library, which can also be easily used within R with the reticulate package. A docker image containing all necessary packages for analysis with an RStudio interface is also available.

## Data Availability

A downsized version of the integrated human atlas containing 10,000 randomly sampled cells is available as a Cerebro (*49*) interactive webpage which can be easily explored by non-bioinformaticians at https://humanlung.iqm.unicamp.br. In Cerebro, users can readily visualize gene expression with dbMAP embeddings, search for each cluster differentially expressed genes, obtain functional enrichment scores for clusters of interest from a wide range of biomedical databases and export publication-level plots and tables. Fully processed data is also available as a complete Seurat object (.Rds) and as a Cerebro (.crb) file. All further data is available from the authors upon request.

## Supplementary Data

A Table with differentially expressed genes (DEG) for each cell type in the lung atlas is available as Lung_markers.csv. Differentially expressed genes (DEG) tables for each cell type in the lung atlas are available as Lung_markers.csv (Supplementary Data).

## Authors contributions

DS-O performed all *in silico* analysis. The original hypothesis of SARS-Cov-2 pathogenesis as an ACE2 dependent impairment on the kinin-kallikrein, renin-angiotensin and coagulation systems was held and explored by LAV, who also coordinated the study and guided exploratory data analysis. The other authors contributed in the interpretation of results and provided suggestions for data analysis. All authors wrote the manuscript and approved the final submitted version.

## Acknowledgments

DS-O received a scholarship from Sao Paulo Research Foundation #2020/04074-2 grant. The study was supported by Sao Paulo Research Foundation grants #2013/08293-7 and #2013/07607-8.

## Supplementary data

Assembly of an integrated human lung cell atlas reveals that SARS-CoV-2 receptor is co-expressed with key elements of the kinin-kallikrein, renin-angiotensin and coagulation systems in alveolar cells

**Supplementary Figure 1.**
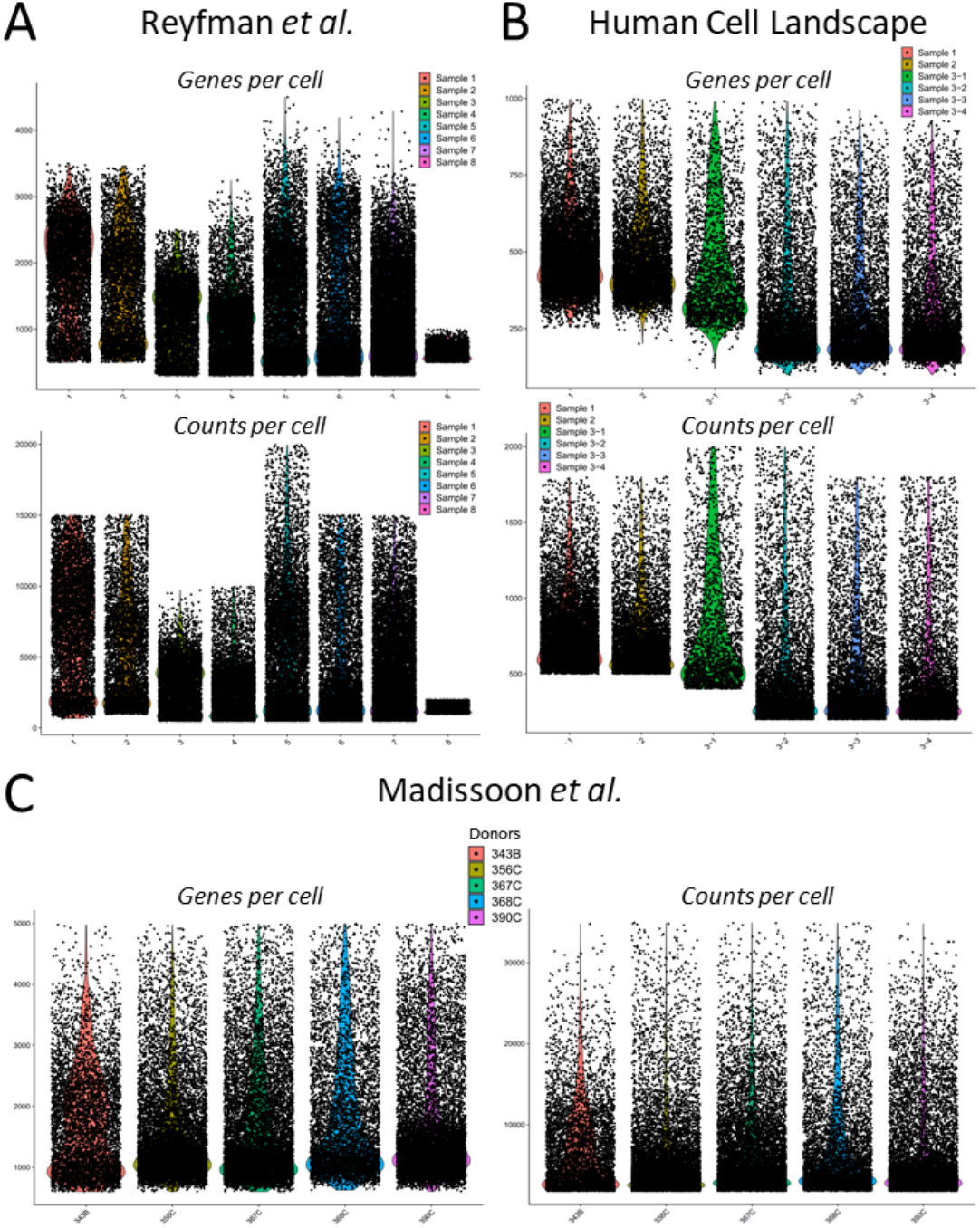
Quality control metrics for included data. Violin plots of quality metrics (detected genes and RNA counts for each cell) showing each of the original datasets used for integration. **a**. Quality control metrics (QC) for Reyfman *et al*. data. **b**. QC for the Human Cell Landscape lung data. **c**. QC for Madissoon *et al*. data.

**Supplementary Figure 2.**
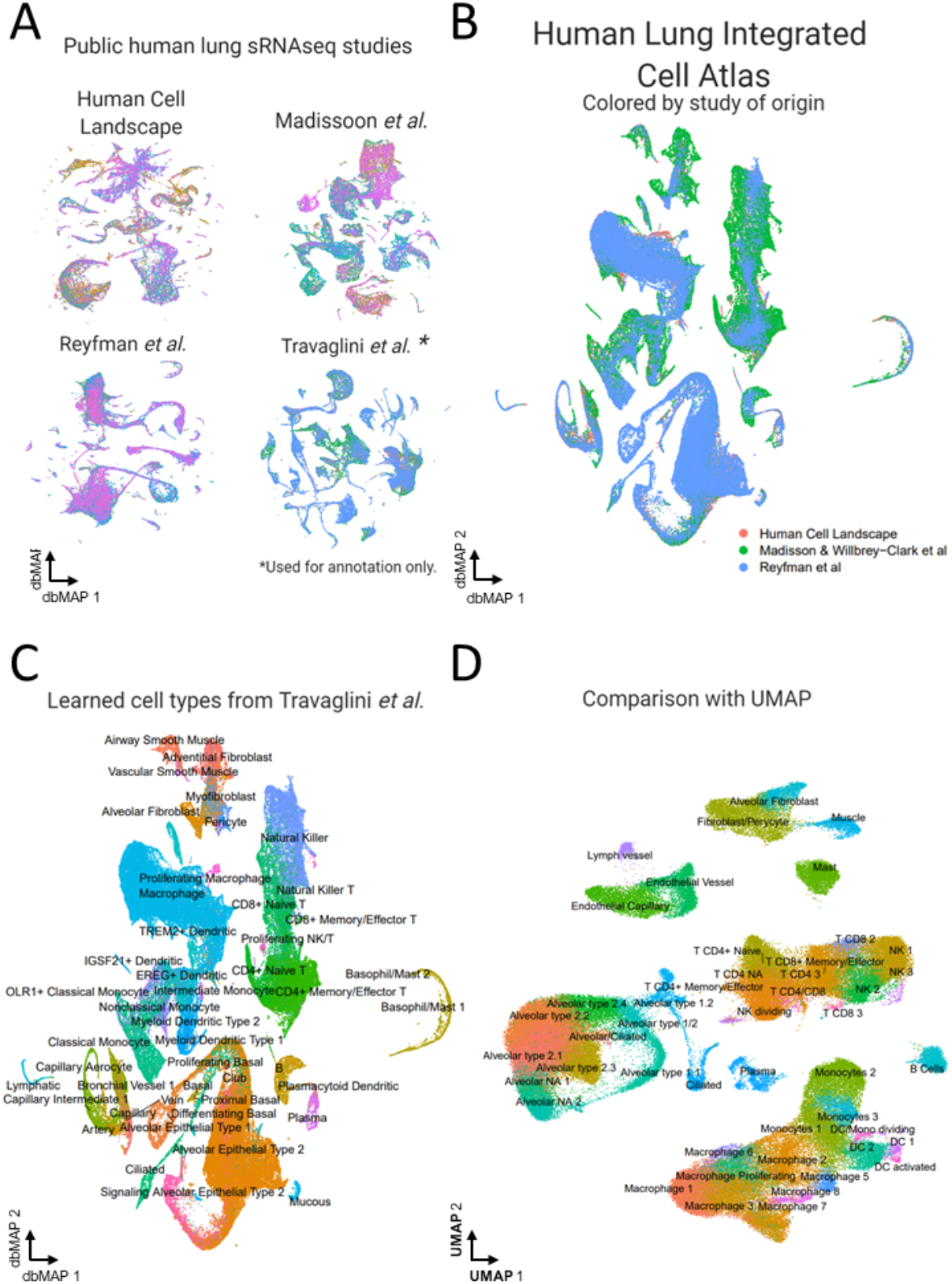
Details on integration, annotation and UMAP layout. **a**. dbMAP embeddings of each individual batch-corrected study prior to integration and label transfer. **b**. dbMAP embedding of the Human Lung Integrated Cell Atlas. Cells were colored by their study of origin. Cell clusters were not particularly enriched for cells from a particular study, and overall integration is able to account for the weighted information from each study. **c**. Labels learned by transfer learning from Travaglini *et al*. annotations. Annotation of resulting clusters and cell-type assignment was partly guided by these annotations. **d**. UMAP layout was computed on top 50 Principal Components after Principal Component Analysis (PCA), the default adopted workflow. Overall cluster configuration is similar between UMAP and dbMAP embeddings, being clearer that dbMAP is advantageous for the visualization of rare populations and differentiation trajectories, taking as example B cells, which are mapped in its differentiation trajectory into plasma cells, whereas UMAP embeds these clusters as completely apart populations. Clusters are annotated by cell type annotation.

**Supplementary Figure 3.**
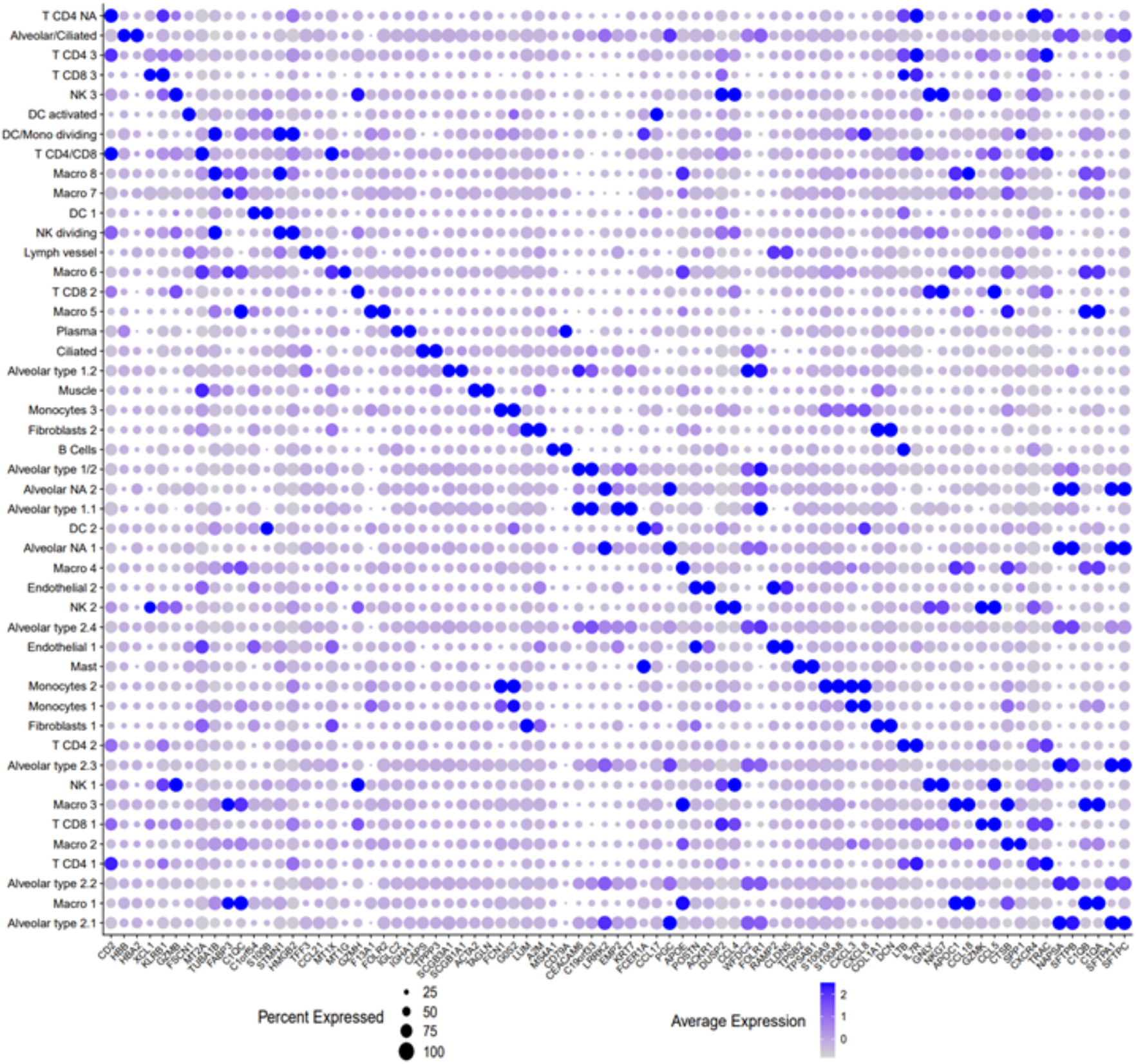
Dotplot of clusters gene expression markers. Dot plot visualization of top 2 highest scoring markers per cell type. Larger circles mean a larger fraction of cells from a specific cell type express that gene, even though at exceptionally low rates. Darker circles mean the average gene expression for that gene in a specific cell-type is higher.

**Supplementary Figure 4.**
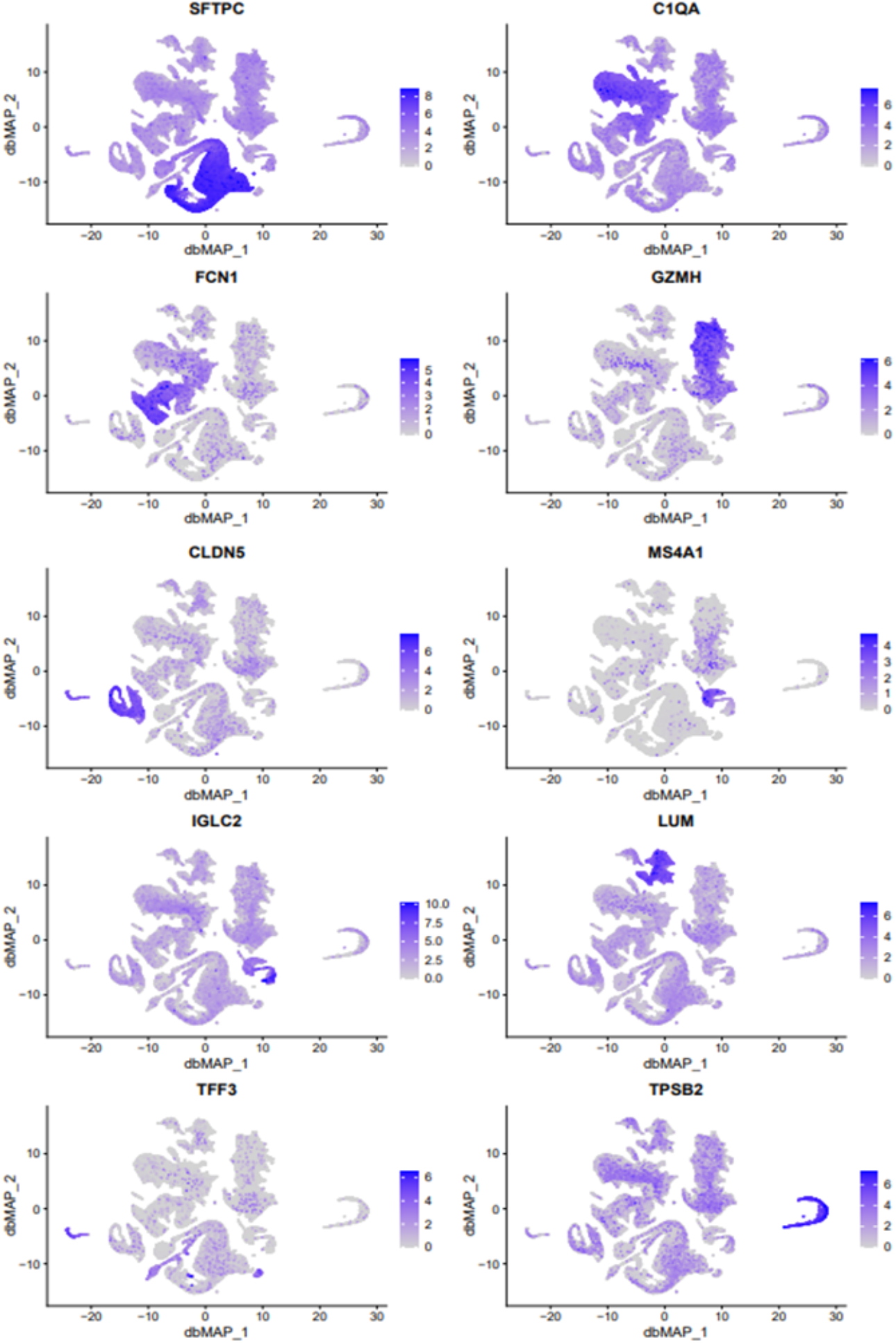
Panel of dbMAP embedded gene expression of 10 cell-type markers. Visualization of gene expression in dbMAP embeddings of the lung atlas. SFTPC: alveolar cells. C1QA: macrophages. FCN1: monocytes. GZMH: T cells. CLDN5: endothelial and lymph vessels cells. MS4A1: B Cells. IGLC2: Plasma cells. LUM: Fibroblasts. TFF3: lymph vessel and alveolar ciliated cells. TPSB2: Mast cells. It is possible to generate similar plots for 20,000 genes on the atlas online database.

**Supplementary Figure 5.**
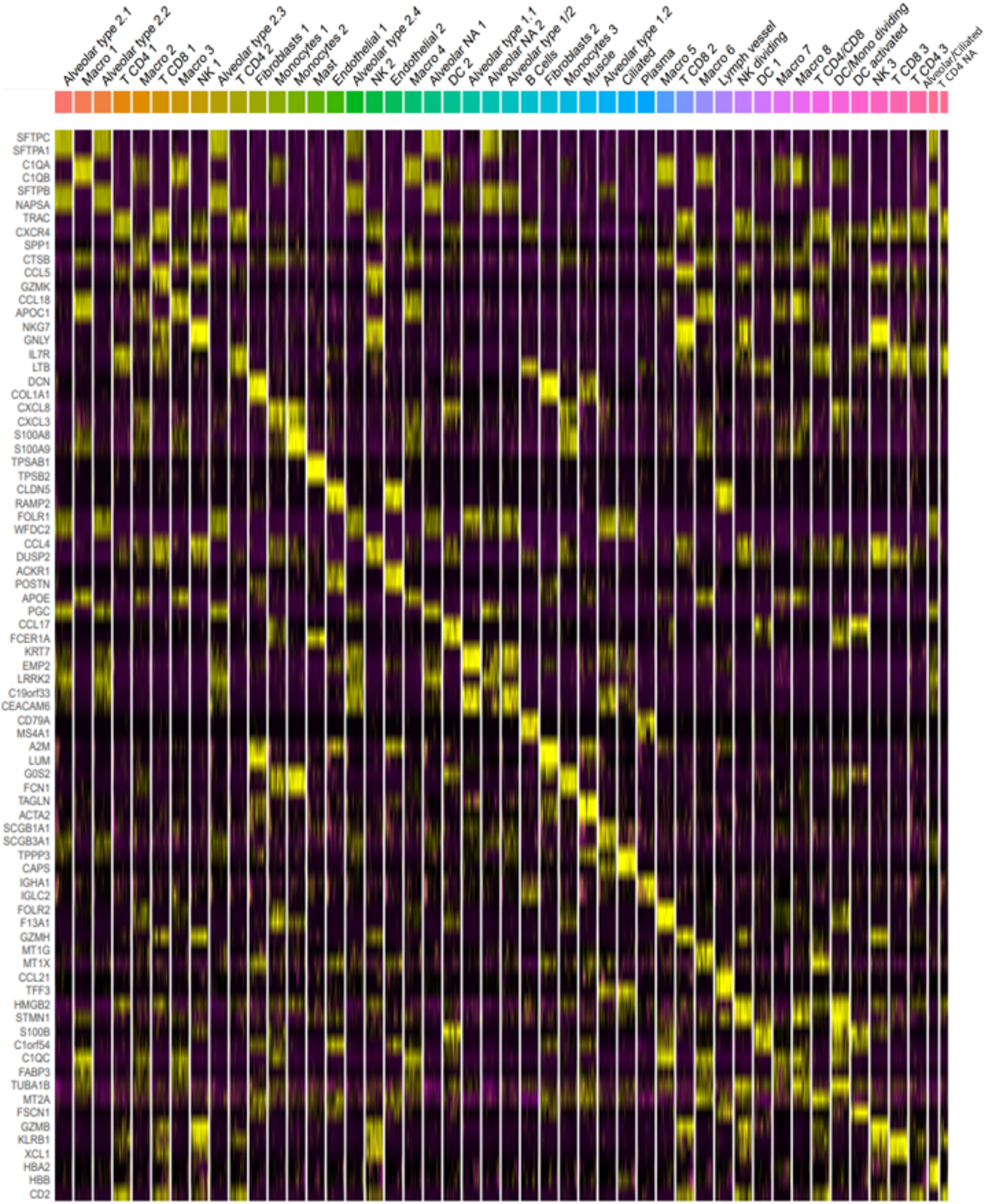
Heatmap of cell-type markers. Heatmap of top two gene expression markers for each cell type cluster. For visualization, the cell number was downsampled by a factor of 100. Top annotations represent cell types.

## Supplementary Tables

**Supplementary Table 1.** Quality control inclusion criteria for cells from analyzed studies. Cells were filtered by a minimum and maximum threshold of detected reads and detected genes so as to avoid overrepresentation of doublets, scRNAseq experimental artifacts that lead to the recognition of multiple cells as one (i.e., two cells in a single droplet). Cells with high mitochondrial gene expression are associated with low-quality reads and were removed from analysis. Each sample from Reyfman *et al*. and Human Cell Landscape data was filtered separately, while Madissoon *et al*. data was of extreme high-quality and presented very low batch-effects, therefore being filtered by a jointly defined threshold. Travaglini *et al*. data was used for annotation purposes only, and also filtered to a jointly defined threshold.

|  | **Reyfman *et al.*** |  |  |  |
| --- | --- | --- | --- | --- |
| Sample | **1** | **2** | **3** | **4** |
| Detected reads | > 500 & < 15,000 | > 1,000 & < 15,000 | > 500 & < 10,000 | > 500 & < 10,000 |
| Detected genes | >500 & < 3,500 | >500 & < 3,500 | > 300 & < 2,500 | > 300 & < 3,500 |
| % Mitochondrial Genes | < 20 | < 10 | < 10 | < 10 |
| Sample | **5** | **6** | **7** | **8** |
| Detected reads | > 500 & < 20,000 | > 500 & < 15,000 | > 500 & < 15,000 | > 1,000 & < 2,000 |
| Detected genes | > 300 & < 4,500 | > 300 & < 5,000 | > 300 & < 4,500 | > 500 & < 1,000 |
| % Mitochondrial Genes | < 20 | < 10 | < 10 | < 11 |
|  | **Madisson *et al.*** |  |  |  |
| Detected reads |  | >1,800 & < 35,000 |  |  |
| Detected genes |  | > 600 & < 5,000 |  |  |
| % Mitochondrial Genes |  | < 10 |  |  |
|  | **Human Cell Landscape** |  |  |  |
| Sample | **1** | **2** | **3** | **4** |
| Detected reads | > 500 & < 1,800 | > 500 & < 1,800 | > 400 & < 2,000 | > 200 & < 2,000 |
| Detected genes | > 250 & < 1,000 | > 200 & < 1,000 | > 100 & < 1,000 | > 100 & < 1,000 |
| % Mitochondrial Genes | < 20 | < 20 | < 20 | < 20 |
| Sample |  | **5** | **6** |  |
| Detected reads |  | > 200 & < 1,800 | > 200 & < 1,800 |  |
| Detected genes |  | > 100 & < 1,000 | > 100 & < 1,000 |  |
| % Mitochondrial Genes |  | < 20 | < 20 |  |
|  | **Travaglini *et al.*** |  |  |  |
| Detected reads |  | > 1,000 & < 40,000 |  |  |
| Detected genes |  | > 600 & < 5,000 |  |  |
| % Mitochondrial Genes |  | < 10 |  |  |

**Supplementary Table 2.** Parameters used for dbMAP embedding for individual studies and the integrated atlas, as well as those used for UMAP embedding of the atlas. dbMAP takes four main parameters. During diffusion, a number **N** of structure components are computed accounting for each cell **K** nearest neighbors. After automatic scaling and selection of relevant components by eigengap analysis, a UMAP layout is generated with **M** as the effective minimum distance between embedded points and **S** as the effective scale of embedded points. Importantly, visualization parametrization can be fine-tuned by the user for its specific dataset due to the fast UMAP layout computation of the structure components, for example by changing the learning rate, although results overall are robust to small changes in these parameters.

|  | Integrated Atlas | Reyfman *et al.* | Madisson *et al.* | Human Cell Landscape |
| --- | --- | --- | --- | --- |
| **UMAP** |  |  |  |  |
| n_PCs | 50 | N/A | N/A | N/A |
| min.dist | 0.5 |  |  |  |
| spread | 1 |  |  |  |
| **dbMAP** |  |  |  |  |
| Computed DCs (N) | 300 | 300 | 200 | 300 |
| Selected DCs (automated) | 191 | 207 | 147 | 169 |
| k-nearest-neighbors (K) | 50 | 50 | 15 | 30 |
| min.dist (M) | 0.3 | 0.3 | 0.6 | 0.3 |
| spread (S) | 2 | 2 | 1 | 1.5 |
| Learning rate | 2 | 1.2 | 1 | 1 |

**Supplementary Table 3.** Coloring thresholds for plots obtained for each gene showed in the analysis. Plots were first visualized with a high threshold, which was decreased as little as possible, so as to visualize gene expression throughout the color scale. The combined thresholds used for the dotplots from figures 3, 4, 5 and 6 are also shown.

| Plotting colorscale thresholds | ACE2 | TMPRSS2 | PIKFYVE | TPCN2 | CTSL | KNG1 | KLKB1 | BDKRB2 | ACE | BDKRB1 |
| --- | --- | --- | --- | --- | --- | --- | --- | --- | --- | --- |
| DimPlots | < 0.1 | < 3 | < 1 | < 0.25 | < 2 | < 0.02 | < 0.1 | < 0.3 | < 0.1 | < 0.06 |
| DotPlot Fig. 3 | None |  |  |  |  |  |  |  |  |  |
| DotPlot Fig. 4 |  |  |  |  |  | < 6 |  |  |  |  |
|  | AGT | REN | AGTR1 | SERPINE1 | PLAT | FGG | SFTPC |  |  |  |
| DimPlots | < 1 | < 1 | < 0.2 | < 0.1 | < 0.3 | < 2 | < 8 |  |  |  |
| DotPlot Fig. 5 | < 3 |  |  |  |  |  |  |  |  |  |
| DotPlot Fig. 6 |  |  |  | < 5 |  |  |  |  |  |  |

**Key Resources Table.** Summary of all data and software used.

| Key Resources Table |  |  |
| --- | --- | --- |
| scRNAseq of human lung | Sequencing technology | Accession |
| Reyfman *et al.* | 10X Genomics v2 | GEO GSE122960 |
|  |  | https://www.ncbi.nlm.nih.gov/geo/query/acc.cgi?acc=GSE122960 |
| Madissoon *et al.* | 10X Genomics v2 | NCBI BIOPROJECT PRJEB31843 |
|  |  | https://www.tissuestabilitycellatlas.org/ |
| Human Cell Landscape | Microwell-seq | GEO GSE134355 |
|  |  | https://www.ncbi.nlm.nih.gov/geo/query/acc.cgi?acc=GSE134355 |
| Softwares and algorithms |  |  |
| R 3.6.2 | R Core | https://www.r-project.org/ |
| Seurat 3.1.5 | Stuart and Butler *et al.* | https://satijalab.org/seurat/ |
| dbMAP v0.1 | Sidarta-Oliveira and Velloso | https://github.com/davisidarta/dbMAP |

